# Global Health Security Index is associated with Covid-19 Pandemic Mortality 2020–2021 but not for Island Jurisdictions

**DOI:** 10.1101/2024.09.02.24312964

**Authors:** Matt Boyd, Michael G Baker, Nick Wilson

## Abstract

**Background:** Past studies show a mixed relationship between the Global Health Security (GHS) Index and Covid-19 pandemic health outcomes. Some recent work that suggested higher GHS Index scores are associated with better mortality outcomes has been criticised on methodological grounds. There remains scope for improved analyses of these relationships, including of island nations and macroeconomic pandemic outcomes.

**Methods:** Multiple linear regression analyses (controlling for per capita GDP and political corruption) across GHS Index scores, age-standardised excess mortality for 2020–2021, and GDP per capita growth, for island and non-island jurisdictions separately.

**Results:** The GHS Index predicted better health outcomes in terms of age-standardised excess mortality through 2020–2021 in non-island jurisdictions (β = -0.046, p = 0.00068, adj R^2^ = 0.45), but not in island jurisdictions (β = 0.027, p = 0.734). For a starting age-standardised excess mortality of 100 per 100 000, a +10-point rise in overall GHS Index score predicts a 26.7 per 100 000 reduction in age-standardised mortality. We found no robust evidence that a higher GHS Index predicted higher year-on-year GDP growth through 2019–2020 or 2020–2021.

**Conclusion:** The GHS Index demonstrated clear associations with favourable health outcomes of non-island jurisdictions through the Covid-19 pandemic, supporting its use to guide pandemic preparedness investments. Contrasting findings for islands suggest the need to enhance how the Index measures border biosecurity capacities and capabilities, including the ability to support the exclusion/elimination strategies that successfully protected islands during the Covid-19 pandemic.

**What is already known?:** The Global Health Security (GHS) Index has been argued to predict excess mortality through the Covid-19 pandemic when accounting for under-reporting and population age-structure.

However, the methodology of some key analyses has been criticised. Furthermore, even if associated with outcomes, it is debated whether the Index is equally applicable across different jurisdictions as a guide to pandemic preparedness.

**What this study adds?:** We analysed 47 island and 142 non-island jurisdictions separately using methodology that addressed published criticism of previous studies. We found that the GHS Index was associated with good health outcomes in non-islands but not in islands. We infer that non-islands and islands exhibited fundamentally different responses and experiences during the Covid-19 pandemic and may require different pandemic policies.

**How might this study affect research, practice, or policy?:** This study confirms the potential of the GHS Index as a starting point for pandemic readiness. Jurisdictions should look to enhance capacities and capabilities most associated with Covid-19 health outcomes. The findings also suggest that border biosecurity, which island states have by virtue of their geography, but other states need to generate by design, may need much greater focus in metrics like the GHS Index.

## INTRODUCTION

### Pandemic threats

The Covid-19 pandemic caused enormous global health and economic impacts. It resulted in the first increase in global all-cause mortality in the 70-year post-war period (a 5.1% increase in the 2020-2021 period compared with a decline of 62.8% from 1950-2019).^1^ Excess mortality during the continuing pandemic period is estimated at 27.3 million deaths up to July 2024.^2^

There is a likelihood of equally or more severe pandemics in the future, including from engineered pathogens.^3-5^ The list of pathogens identified with pandemic potential is growing.^6^ Several are causing current concern, notably the influenza A(H5N1) panzootic now infecting cattle in the US.^7^ There is also a more harmful Mpox clade currently spreading in Africa that has become the eighth public health emergency of international concern to be declared by WHO.^8^ These actual and potential harms highlight the benefits from implementing effective preparedness measures (as described in extensive national pandemic reviews such as those conducted in the UK,^9^ Australia,^10^ and New Zealand^11^).

### GHS Index and Covid-19 pandemic

The Global Health Security (GHS) Index is a comprehensive, criteria-based assessment of health security capabilities across 195 States Parties to the International Health Regulations. The Index was first published in 2019 by the Nuclear Threat Initiative (NTI), Johns Hopkins Center for Health Security, and the Economist Intelligence Unit.^12^ The metric encompasses six categories (that can be abbreviated as ‘Prevention’, ‘Detection and Reporting’, ‘Rapid Response’, ‘Health System’, ‘Compliance with International Norms’, and ‘Risk Environment’), which are composed of 37 indicators and various sub-indicators. It quantifies countries’ abilities or potential to carry out public health functions necessary for infectious disease outbreak prevention, detection and response, by giving an ‘Overall Score’ out of 100.

The GHS Index has face validity,^13^ and has been validated against communicable disease mortality globally with higher scores predicting a lower burden of communicable disease deaths by country.^14^ On this basis one would expect countries with higher GHS Index scores to have had better health outcomes through the Covid-19 pandemic.

However, studies of the relationship between Covid-19 outcomes and GHS Index scores early in the Covid-19 pandemic did not reveal the expected relationships. Paradoxically, high GHS Index scores (better prepared countries) looked to have worse Covid-19 outcomes in terms of Covid-19 deaths per capita and number of cases,^15-18^ as well as detection and response times,^19^ or the GHS Index had ‘no explanatory power’^20^ and pandemic preparedness indices generally were, ‘not meaningfully associated with standardised infection rates or infection fatality ratios.’^21^ Furthermore, Covid-19 outcomes were significantly associated with sociodemographic, political and governance variables not included in the 2019 GHS Index. Social cohesion, reduction in social polarisation and reduced perceptions of corruption were consistently correlated with lower excess mortality associated with the pandemic.^22^ Finally, the GHS Index did not sufficiently consider the importance of geography, with island nations such as Australia, New Zealand, and Pacific islands able to close their borders to prevent the virus from entering.^23^

On the other hand, some studies have shown the expected correlations between GHS Index scores and pandemic outcomes. For example, higher GHS Index scores were associated with low R0 values across 52 countries.^24^ African countries that fell in the ‘more prepared’ GHS Index category had fewer deaths and cases of Covid-19 than those in the ‘least prepared’ category.^25^ When deaths per capita later in the pandemic were analysed (July 2021), there was a strong negative correlation (r = -0.69, p < 0.01) between European countries’ GHS Index scores and their excess mortality rates, indicating that countries with higher GHS Index scores had lower excess mortality during the pandemic.^26^ Another study found a statistically significant slight negative correlation between GHS Index scores and excess mortality at the global scale.^27^

With respect to age, for each percentage point increase in the proportion of the population over age 65 years, excess mortality at 100 days in the Covid-19 pandemic increased by 3.35 deaths per 100 000 individuals (p=0.006).^22^ The 2023 study by Ledesma et al. accounted for age-standardised excess mortality (n = 183 countries) and demonstrated an association between higher GHS Index scores and fewer Covid-19 deaths globally. Each 5-point increase in the GHS Index was associated with a 0.21 lower Comparative Mortality Ratio (CMR) for excess Covid-19 mortality, after adjusting for GDP per capita.^28^ This latter study highlighted several problems with some of the previous analyses including with data quality and comparability, and the fact that different country population structures means that age-standardised mortality data is needed. However, it was itself criticised for choices of statistical thresholds, and for not transforming the skewed GDP data used as a control variable.^29^

### Islands and the Covid-19 pandemic

The inclusion of island nations likely skews many analyses of the relationship between GHS Index and Covid-19 pandemic outcomes. This is because island nations, especially small developing island nations in the Pacific, exhibit very low GHS Index scores.^13^ ^30^ However, being an island country was associated with lower excess mortality during the pandemic.^22^ Islands singled out as performing particularly well include New Zealand, Australia, Singapore, Iceland, Taiwan, and the pseudo-island of South Korea.^17^ ^31^ Also, the Oceania region, which is primarily composed of island nations, was noted to have the lowest excess mortality rate of any region.^27^

A likely explanation for the relative success of island jurisdictions is that they are surrounded by ocean, and many implemented effective border controls during the pandemic, potentially rendering irrelevant many internal factors that might contribute to GHS Index scores. Being an island also significantly increased the probability of a country pursuing a Covid-19 elimination strategy.^20^ Indeed ‘island status’ (reduced Covid-19 mortality) along with ‘aged population’ (increased mortality) were among the factors most correlated with excess mortality in one analysis.^22^

### GDP and the Covid-19 pandemic

Potentially confounding any relationship between GHS Index scores and excess mortality is the fact that per capita gross domestic product (GDP) was a good predictor of excess mortality in 20 European countries (r = −0.71, p < 0.01).^26^ Additionally, GDP has been previously correlated with GHS Index scores,^13^ ^14^ and excess mortality is broadly, though not consistently, associated with lower sociodemographic index (SDI) scores.^1^

A number of studies have found that countries using a strong proactive response to the Covid-19 pandemic or using an elimination strategy, resulted in more favourable GDP growth outcomes when compared to countries adopting a mitigation/suppression strategy.^20^ ^32-34^ But none of these studies examined any relationship between pre-pandemic planning/GHS Index with Covid-19 pandemic macroeconomic outcomes.

### Aims and hypotheses

All these findings suggest that a more definitive analysis of the relationship between GHS Index and Covid-19 outcomes should focus on excess mortality rather than number of deaths, age-standardise the health data, explore outcomes other than just death, such as economic impacts, appropriately control for GDP per capita, and analyse islands and non-islands separately. We sought to explore the associations among these variables.

Specifically, we sought to address some of the methodological shortcomings of previous attempts to analyse the association between GHS Index scores and Covid-19 pandemic outcomes, by exploring the following research questions:

1. Whether any relationships between GHS Index scores and excess mortality due to Covid-19 remain after appropriate data transformations and statistical tests.
2. Furthermore, whether there are any associations between GHS Index scores and macroeconomic performance through the pandemic period.
3. Whether consideration of island and non-island jurisdictions separately demonstrates any important patterns in relationships between GHS Index scores and the above Covid-19 pandemic health and/or macroeconomic outcomes.

## METHODS

### Study jurisdictions

Our dataset included 204 jurisdictions with excess mortality data for the first two years of the Covid-19 pandemic. We divided the dataset into two groups, ‘island’ jurisdictions and ‘non-island’ jurisdictions. The study population comprised all jurisdictions within these groups that had previously been scored according to the GHS Index.^13^

We defined island jurisdictions as those surrounded by water, while ignoring structural connections to other land masses such as bridges and tunnels (and so including Singapore and the UK as islands). All these included island jurisdictions also had to have GHS Index data, which excluded sub-national islands (eg, Hawaii). In summary, the group of island jurisdictions included the following: individual sovereign islands, island archipelagos (eg, Indonesia, Philippines), island continents (ie, Australia), island jurisdictions that were non-sovereign but had GHS Index data eg, Cook Islands (New Zealand), and where the jurisdiction had a land border with another jurisdiction on the same island (eg, Ireland, UK, Timor-Leste, Papua New Guinea, Brunei, Dominican Republic, Haiti, Cyprus). We excluded South Korea (although it can be considered a pseudo-island) and jurisdictions with mixed characteristics but where the capital city was on a continental land mass (eg, Malaysia, Corsica [France], Sardinia and Sicily [Italy]).

### Data sources

#### GHS Index scores

We obtained data on jurisdiction-level capacities and preparedness against biological threats from the pre-Covid-19 pandemic (2019) and latest (2021) versions of the GHS Index. Data on the overall score, six category scores, and 37 indicator scores were extracted for analyses.^12^ ^13^

#### Covid-19 health outcomes

We obtained excess mortality and age-standardised excess mortality by year, for these jurisdictions, for 2020 and 2021 from the GBD Study Demographics Collaborators. The estimation methods for these data have previously been described in detail.^1^ Excess mortality is an important measure of the true mortality impact from the Covid-19 pandemic because it identifies the difference between observed all-cause mortality and mortality expected under normal conditions. This dataset demonstrates approximately 16 million global deaths due to Covid-19, with 20 jurisdictions having negative excess mortality in either 2020 or 2021.

#### Macroeconomic outcomes

We obtained GDP data for these jurisdictions from the World Bank’s World Development Indicators, namely GDP per capita purchasing power parity (PPP) (constant 2017 international $), which included adjustment for inflation, ie, the World Bank dataset “NY.GDP.PCAP.PP.KD”, and calculated GDP growth from 2019 to 2020, and from 2020 to 2021, based on 5-year geometric means.

#### Control variables

Our primary control variable was mean GDP PPP per capita (as above) for the 5-year period ending in 2019. We also obtained data from the Covid-19 NP Collaborators for the three variables they found consistently associated with Covid-19 outcomes.^21^ These data represented the level of government corruption, trust in individuals, and trust in government. The data had been developed through principal components analysis of underlying global survey data, as described by the Covid-19 NP Collaborators in 2022.^21^ However, with the exception of government corruption, data was missing for the majority of island jurisdictions limiting potential inclusion.

#### Missing data

Not every jurisdiction had data available for each variable. Additionally, the coverage gaps were not the same for each variable. We ignored, rather than attempted to estimate this missing data. This meant in practice that N varied across our analyses, and there was rapid reduction in N with inclusion of additional control variables beyond GDP.

In the main analysis, the following jurisdictions were excluded as we had GHS Index scores, but not GDP PPP from the World Bank dataset: Bahamas, Cook Islands, Cuba, Niue, Andorra, Eritrea, Monaco, North Korea, South Sudan, Syria, Venezuela, and Yemen. For all results we provide the number of jurisdictions (both island and non-island), which were included.

### Analyses performed

We conducted both epidemiological and macroeconomic analyses and took a hierarchical approach to the GHS Index, it’s six categories and the indicators that comprise the categories. Due to right-skewness we log-transformed GDP per capita by taking the natural logarithm, as is common in econometric analysis.^29^ Similarly, we transformed age-standardised excess mortality by taking the cube root, since natural logarithms were not possible due to negative values.

First, we conducted multiple linear regression taking the GHS Index overall score (level 1 of the GHS Index) as the independent variable. We examined, in turn, the dependent variables of age-standardised excess mortality, GDP per capita growth 2019 to 2020, and GDP per capita growth 2020 to 2021, for each group: all jurisdictions, islands, and non-islands.

With the aim of demonstrating the impact of including variables as in previously published work versus correcting for methodological weaknesses, we examined three regression models in turn plus a sensitivity analysis. First (Model 1), we replicated the 2023 analysis of Ledesma et al.^35^ by conducting regressions using 2021 GHS Index scores, with untransformed GDP and excess mortality variables, while controlling for GDP per capita (in current $). Second (Model 2), we conducted the same analyses using 2019 (pre-pandemic) GHS Index scores, using both GDP (PPP) and excess mortality data transformed as above.

Third (Model 3 – designated our main analysis), we included the additional control variable representing government corruption. We performed exploratory analysis with the addition of the other two control variables (trust in government and trust in individuals), understanding that this would greatly reduce the number of jurisdictions able to be studied. We provide the regression equations for these analyses in the Supplementary Material.

Each primary analysis of GHS Index overall score involved nine comparisons (within each model) and we applied the corresponding Bonferroni corrected p-value threshold for statistical significance (0.0056).

For statistically significant associations in the main analysis (following Bonferroni correction), we then extracted regression results across the six category scores (level 2) as independent variables, and if applicable, the indicator scores (level 3) comprising those categories. At the category and indicator level we adjusted for multiple analyses using the Benjamini-Hochberg correction with a false discovery rate threshold of 0.05. This is because the Bonferroni may be overly conservative when there is a hypothesised relationship between the dependent and independent variables, and when the independent variables are correlated (as they are across the category scores).

We do not report pure correlation results (Pearson’s r or Spearman’s rho) as the existence of these statistically significant relationships has been established in previous work,^35^ and further reporting them without controlling for key variables such as GDP per capita risks distracting from important controlled findings.

## RESULTS

Descriptive statistics across jurisdictional groupings (all jurisdictions, islands, non-islands) are presented in Table 1. Supplementary Table S1 shows data coverage (number of jurisdictions with data) across each variable. Jurisdiction-specific age-standardised excess mortality for 2020–2021 across all locations ranged from -59.6 to 897.4 per 100 000 population (n = 204). Islands demonstrated lower excess mortality (mean 59 vs 193 for non-islands). Mean GDP per capita growth was 0.38% from 2019 to 2020, and 0.8% from 2020 to 2021 across all jurisdictions (n = 184, World Bank GDP [PPP] data). The mean 2021 GHS Index score of non-islands was 38.9 (n = 145) and islands 35.0 (n = 49) out of 100.

**Table 1:**
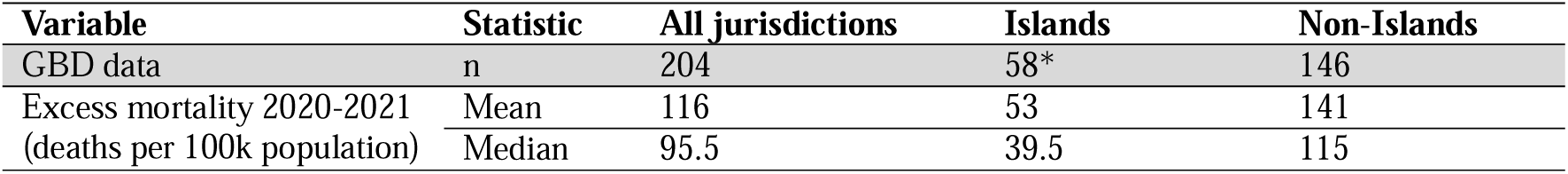
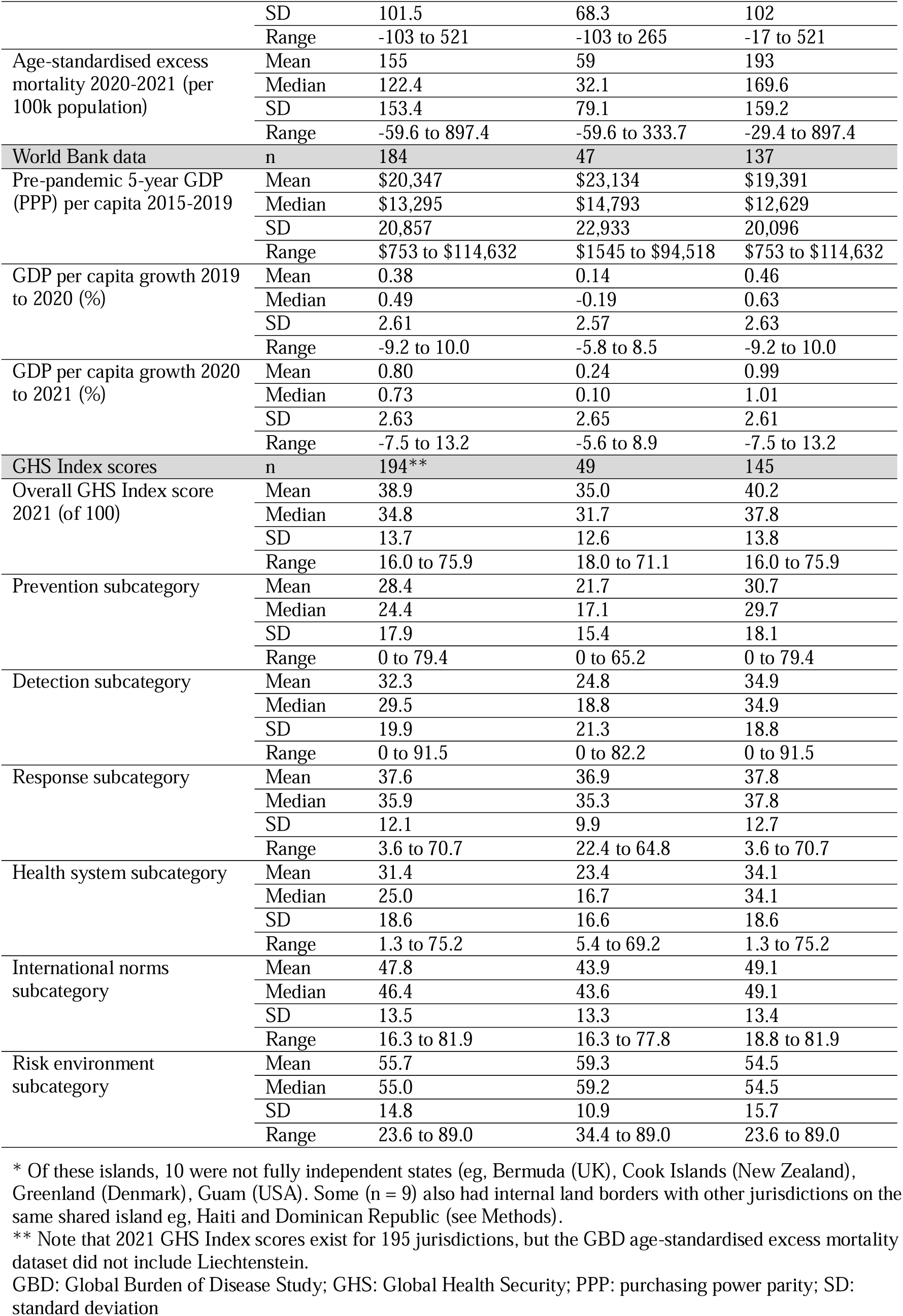
Descriptive values for jurisdiction groups included in the analysis

### Multivariable analyses

Results of the primary regression analyses comparing GHS Index overall score with the three outcome variables, across the three analysis models are presented in Table 2. Distributions across key variables, pre- and post-transformation, are shown in Supplementary Figures S1 and S2.

**Table 2:** First-level of hierarchical analysis (GHS Index Overall Score vs 3 dependent variables): Regression results across all 3 Models with significant P-values after Bonferroni correction asterisked for each of the three analysis families

| Group | Model | Depend. Variable | Indep. Variable | Control Variables | Coefficient (SE)<sup>#</sup> | T-Value | P-Value | R<sup>2</sup> | Adj. R<sup>2</sup> | N |
| --- | --- | --- | --- | --- | --- | --- | --- | --- | --- | --- |
| **Age-standardised excess mortality 2020–2021** |  |  |  |  |  |  |  |  |  |  |
| All | 1 | EM | GHSI 2021 | GDP | -3.091 (0.86) | -3.61 | 0.000387* | 0.22 | 0.21 | 189 |
| All | 2 | $\sqrt[3]{EM}$ | GHSI 2019 | ln(GDP) | 0.011 (0.01) | 0.79 | 0.432050 | 0.20 | 0.19 | 182 |
| All | 3 | $\sqrt[3]{EM}$ | GHSI 2019 | ln(GDP)+ corruption | -0.005 (0.01) | -0.38 | 0.704835 | 0.46 | 0.45 | 155 |
| Islands | 1 | EM | GHSI 2021 | GDP | 0.647 (1.53) | 0.42 | 0.675088 | 0.10 | 0.06 | 47 |
| Islands | 2 | $\sqrt[3]{EM}$ | GHSI 2019 | ln(GDP) | 0.027 (0.04) | 0.75 | 0.458671 | 0.06 | 0.01 | 45 |
| Islands | 3 | $\sqrt[3]{EM}$ | GHSI 2019 | ln(GDP)+ corruption | 0.012 (0.03) | 0.34 | 0.733720 | 0.53 | 0.46 | 27 |
| Non-Islands | 1 | EM | GHSI 2021 | GDP | -5.353 (0.89) | -5.99 | <0.000001* | 0.36 | 0.35 | 142 |
| Non-Islands | 2 | $\sqrt[3]{EM}$ | GHSI 2019 | ln(GDP) | -0.050 (0.01) | -4.46 | 0.000017* | 0.46 | 0.45 | 137 |
| Non-Islands | 3 | $\sqrt[3]{EM}$ | GHSI | ln(GDP)+ | -0.046 (0.01) | -3.48 | **0.000683\*** | 0.49 | 0.48 | 128 |

| Islands |  |  | 2019 | corruption |  |  |  |  |  |  |
| --- | --- | --- | --- | --- | --- | --- | --- | --- | --- | --- |
| GDP Growth 2019-2020 |  |  |  |  |  |  |  |  |  |  |
| All | 1 | GDP Growth | GHSI 2021 | GDP | 0.076 (0.03) | 2.71 | 0.007405 | 0.04 | 0.03 | 188 |
| All | 2 | GDP Growth | GHSI 2019 | ln(GDP) | 0.057 (0.02) | 3.36 | 0.000951* | 0.07 | 0.06 | 182 |
| All | 3 | GDP Growth | GHSI 2019 | ln(GDP)+ corruption | 0.061 (0.02) | 2.89 | **0.004457\*** | 0.09 | 0.07 | 155 |
| Islands | 1 | GDP Growth | GHSI 2021 | GDP | -0.047 (0.07) | -0.71 | 0.479910 | 0.01 | -0.03 | 47 |
| Islands | 2 | GDP Growth | GHSI 2019 | ln(GDP) | 0.030 (0.04) | 0.84 | 0.403055 | 0.03 | -0.02 | 45 |
| Islands | 3 | GDP Growth | GHSI 2019 | ln(GDP)+ corruption | 0.048 (0.05) | 0.95 | 0.351436 | 0.05 | -0.08 | 27 |
| Non-Islands | 1 | GDP Growth | GHSI 2021 | GDP | 0.106 (0.03) | 3.23 | 0.001563* | 0.07 | 0.06 | 141 |
| Non-Islands | 2 | GDP Growth | GHSI 2019 | ln(GDP) | 0.074 (0.02) | 3.46 | 0.000719* | 0.09 | 0.08 | 137 |
| Non-Islands | 3 | GDP Growth | GHSI 2019 | ln(GDP)+ corruption | 0.060 (0.03) | 2.36 | 0.019763 | 0.11 | 0.08 | 128 |
| GDP Growth 2020-2021 |  |  |  |  |  |  |  |  |  |  |
| All | 1 | GDP Growth | GHSI 2021 | GDP | 0.084 (0.03) | 3.29 | 0.001202* | 0.07 | 0.06 | 187 |
| All | 2 | GDP Growth | GHSI 2019 | ln(GDP) | 0.060 (0.02) | 3.47 | 0.000649* | 0.07 | 0.06 | 181 |
| All | 3 | GDP Growth | GHSI 2019 | ln(GDP)+ corruption | 0.053 (0.02) | 2.47 | 0.014449 | 0.06 | 0.04 | 155 |
| Islands | 1 | GDP Growth | GHSI 2021 | GDP | 0.018 (0.07) | 0.24 | 0.810317 | 0.02 | -0.03 | 46 |
| Islands | 2 | GDP Growth | GHSI 2019 | ln(GDP) | 0.051 (0.04) | 1.41 | 0.166045 | 0.05 | 0.01 | 45 |
| Islands | 3 | GDP Growth | GHSI 2019 | ln(GDP)+ corruption | 0.044 (0.05) | 0.89 | 0.382546 | 0.08 | -0.04 | 27 |
| Non-Islands | 1 | GDP Growth | GHSI 2021 | GDP | 0.093 (0.03) | 3.20 | 0.001728* | 0.08 | 0.07 | 141 |
| Non-Islands | 2 | GDP Growth | GHSI 2019 | ln(GDP) | 0.059 (0.02) | 2.72 | 0.007440 | 0.06 | 0.04 | 136 |
| Non-Islands | 3 | GDP Growth | GHSI 2019 | ln(GDP)+ corruption | 0.049 (0.03) | 1.89 | 0.060968 | 0.07 | 0.05 | 128 |

**Table notes:** Asterisk (\*) indicates p-value remains statistically significant against the pre-specified Bonferroni corrected p threshold of 0.0056; $\sqrt[3]{\cdot}$ : cube root transformed; Corruption: government corruption; EM: age-standardised excess mortality; GHSI: Global Health Security Index; ln: natural logarithm transformed; SE: standard error.

In Model 1, similar to the findings of previous published analyses, without transformations of mortality and GDP variables, and using current value GDP,^35^ there is an association between higher 2021 GHS Index overall scores and lower age-standardised excess mortality during 2020-2021, across all jurisdictions (β = -3.091 [SE = 0.855, 95% CI: -4.78 to -1.40], F = 26.6, p = 0.000387), controlling for GDP per capita. This association is stronger in non-islands (β = -5.353 [SE = 0.893, 95% CI: -7.12 to -3.59], F = 38.8, p < 0.000001), explaining 35% of the variance in mortality rates across 142 jurisdictions, but the association was not found in island jurisdictions. However, as noted in critiques of previous studies,^29^ this statistical association disappears for the ‘all jurisdictions’ group when pre-pandemic 2019 GHS Index scores, log-transformed GDP (PPP), and cube root transformed age-standardised excess mortality are used (Model 2).

Nevertheless, when analysing islands and non-island separately, the association persists in Model 2 for non-islands (β = -0.050, [SE = 0.011, 95% CI: -0.073 to -0.028], F = 57.4, p = 0.000017; note β in Model 2 cannot be directly compared to Model 1 due to cube root transformation of excess mortality in Model 2). When control for government corruption is added in Model 3, there remains an association between 2019 GHS Index overall score and age-standardised excess mortality for non-island jurisdictions (β = -0.046, [SE = 0.013, 95% CI: -0.071 to -0.020], F = 39.8, p = 0.000683, n = 128, explaining 48% of variance). The p-value is lower than our Bonferroni corrected threshold of 0.0056 (9 comparisons in this primary analysis) and it is lower than the threshold of 0.0009 used by Ledesma et al. across their 57 hypotheses.

Figure 1 illustrates the relationship between 2019 GHS Index overall score and predicted age-standardised excess mortality (Model 3, controlling for GDP [PPP] and government corruption). The relationship is non-linear due to the cube root transformation. Figure 1 shows, for example, that from a starting excess mortality of 100 per 100 000, a +10-point shift in overall GHS Index score is associated with -26.7 per 100 000 mortality reduction, equating to 2670 lives saved in a hypothetical country of 10 million population. Figure 2 depicts the regression line and 95% CI for GHS Index overall score (2019) vs age-standardised excess mortality 2020–2021 (Model 3), for all jurisdictions, islands and non-islands.

**Figure 1:**
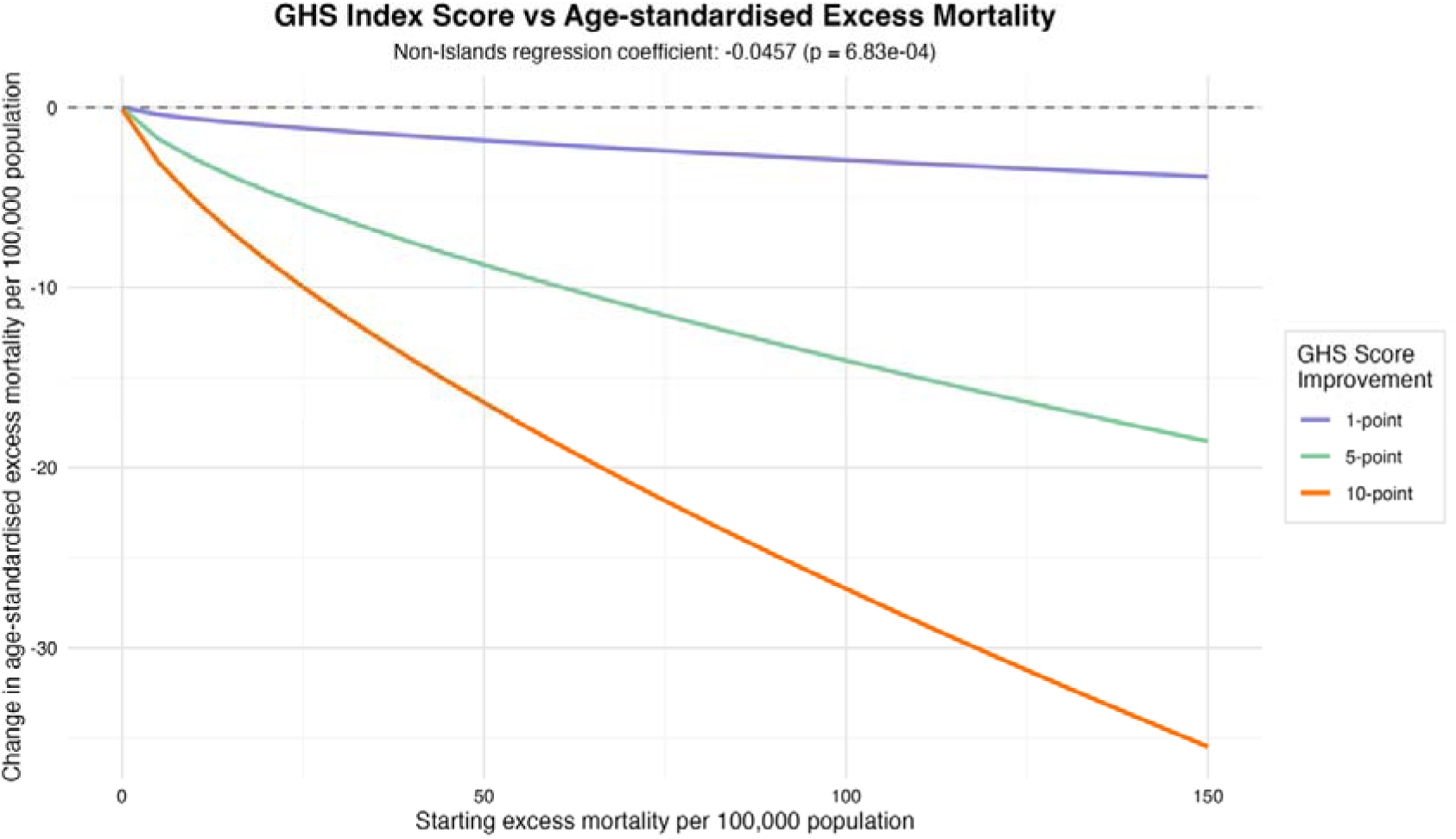
Effect of 2019 GHS Index overall score on age-standardised excess mortality 2020–2021 for non-island jurisdictions based on Model 3, controlling for GDP (PPP) per capita and government corruption (non-linear effect results from cube root transformation of excess mortality data)

**Figure 2:**
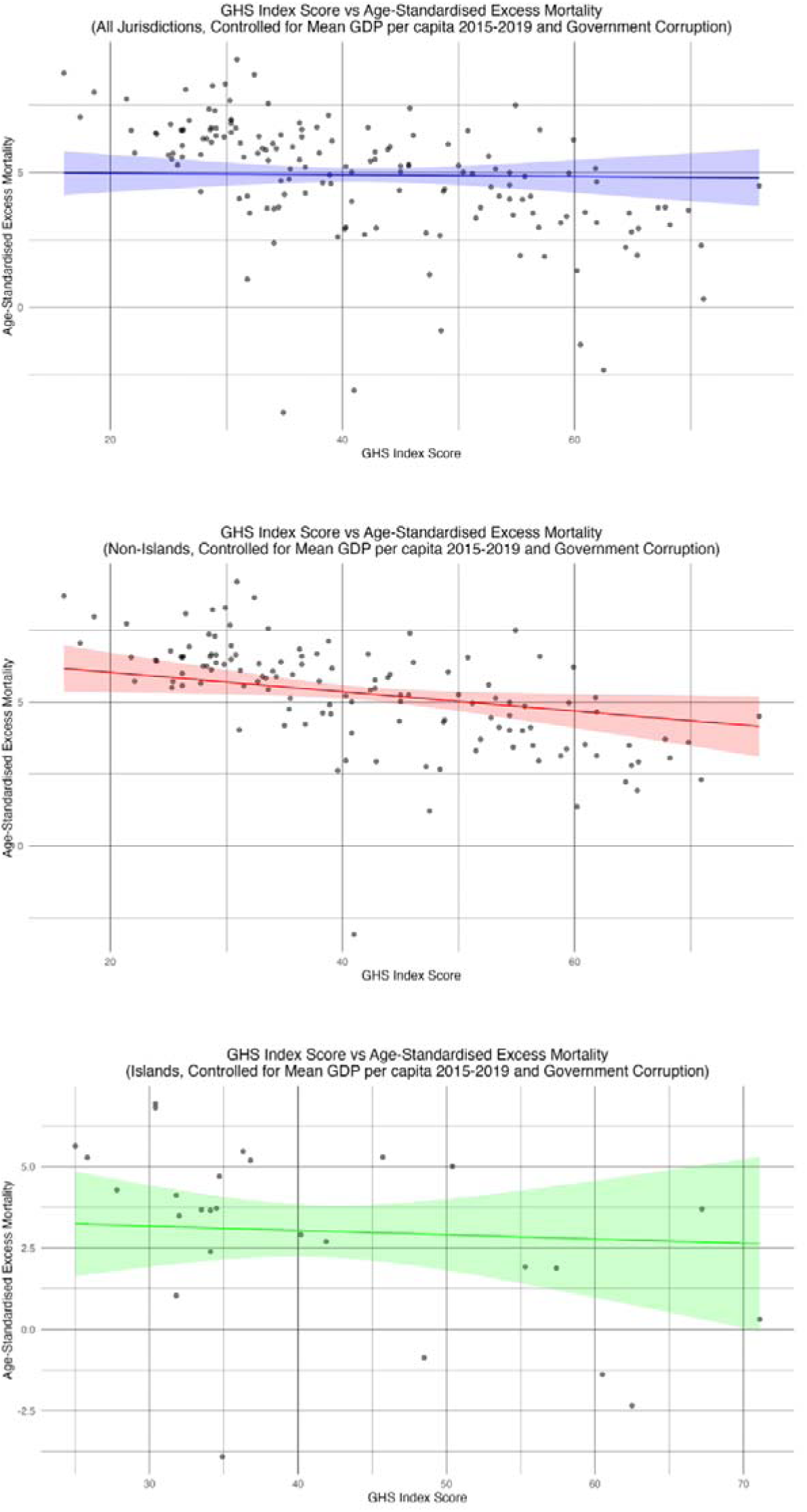
Relationships (Model 3) between the 2019 Global Health Security Index overall score and ∛(age-standardised excess mortality) 2020–2021 for all jurisdictions, non-islands, and islands. The black points represent jurisdictions while the coloured lines represent linear regression lines for the relationships with shaded areas representing corresponding 95% CIs.

For the dependent variable of GDP growth, a statistical association with p value below our Bonferroni corrected threshold persists in our main analysis Model 3 only for the ‘all jurisdictions’ group for GDP growth in the first year of the pandemic (Table 2).

Based on the foregoing results, we took the GHS Index category scores (level 2 of the GHS Index) for non-islands forward for regression analysis (Model 3) against the dependent variable of age-standardised excess mortality, and for all jurisdictions against GDP growth 2019 to 2020.

Table 3 presents results of the second-level of our hierarchical analysis. After adjustment for both mean GDP (PPP) per capita 2015–2019 and government corruption. In this analysis p values are corrected using the Benjamini-Hochberg method, as we already hypothesise relationships (based on our level-one analysis) and the independent variables are known to be correlated. Results show that all 2019 GHS Index categories are associated with 2020–2021 age-standardised excess mortality in non-islands, except for the ‘compliance with international norms’ category. For GDP growth 2019-2020, although Table 3 shows some statistical associations, model fit based on r-squared statistic is universally very poor, and so we should not read much into these results.

**Table 3:**
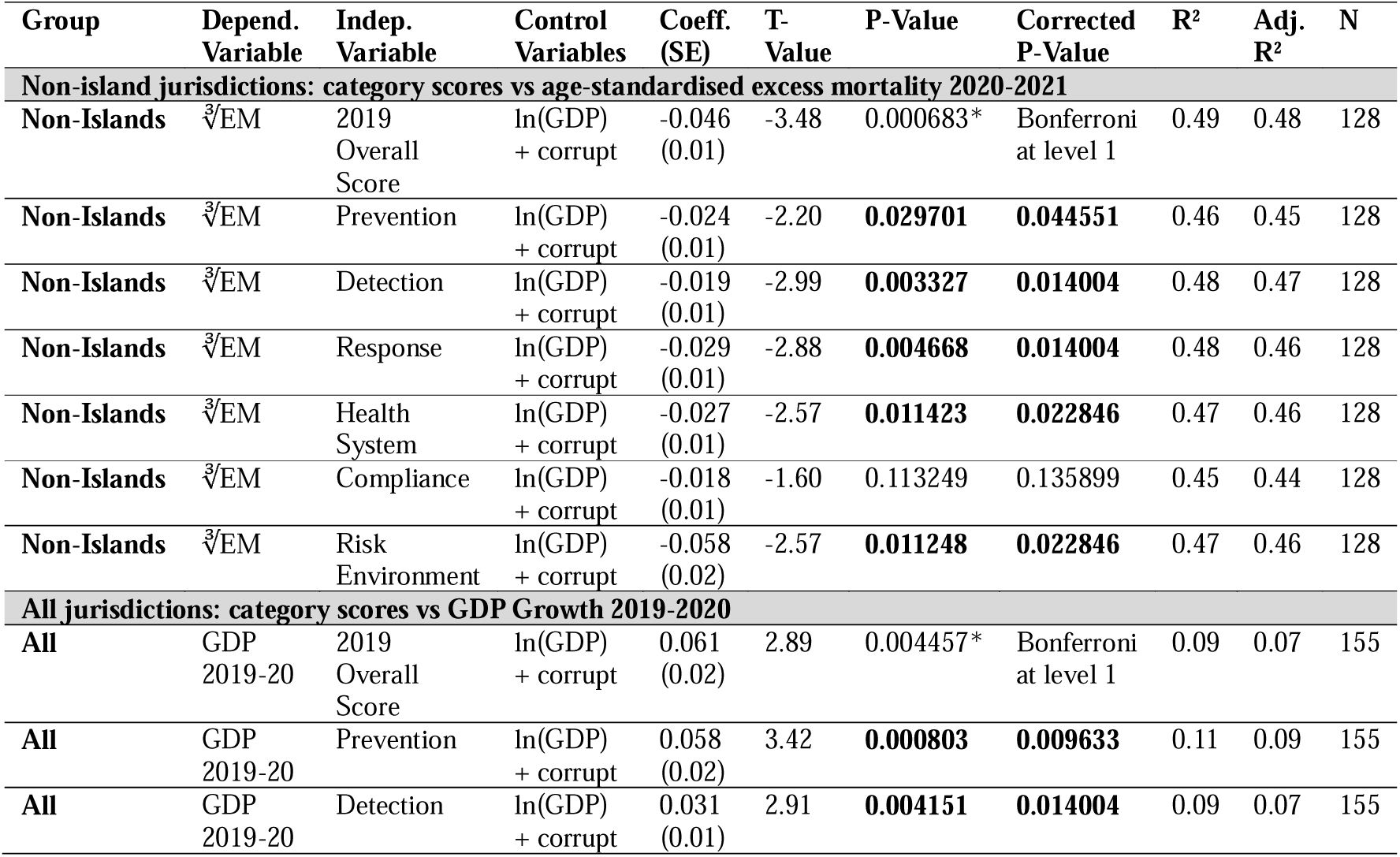
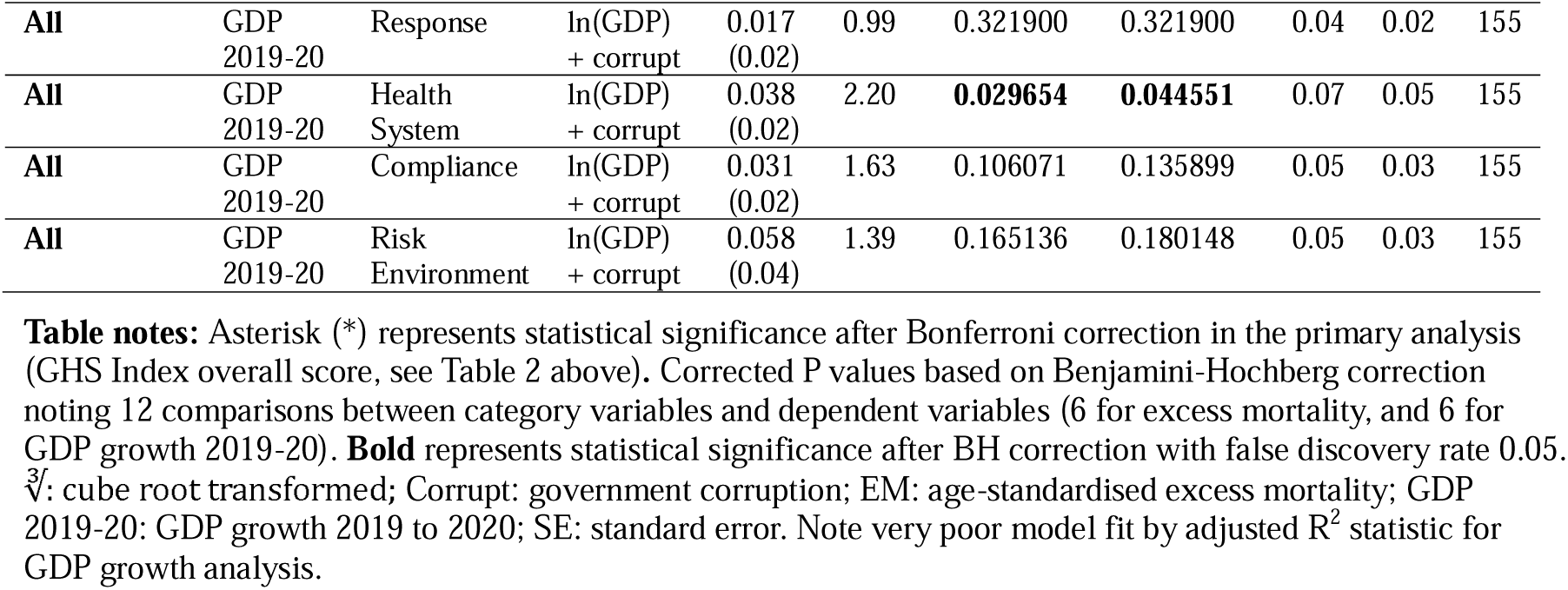
Second-level hierarchical analysis (2019 GHS Index Category Scores): Regression results (Model 3) for those jurisdictions and dependent variables showing statistical significance at first-level analysis

Following inspection of histogram distributions of the data underlying the 2019 GHS Index indicator level scores (see Supplementary Material Table S3a-d) we chose not to pursue a ‘level three’ analysis of the GHS Index because it is clear the data at this level (unlike overall score and category scores) strongly violate the assumptions of linear regression.

### Sensitivity analysis

Supplementary Tables S2 to S4 present the full ‘overall score’ regression results including an exploratory sensitivity analysis additionally controlling for trust in government and trust in individuals, as well as GDP and government corruption. Supplementary Tables S5 to S7 present full exploratory regressions across all category scores for all jurisdictions.

The sensitivity analysis including all control variables suggests that the statistical relationship between excess mortality and non-island GHS Index overall score holds (β = -0.052, [SE = 0.017, 95% CI: -0.085 to -0.019], F = 24.5, T-value -3.14, p = 0.002844), with the model explaining 68% of variance in excess mortality. There were no statistically significant associations with GDP growth. However, only 56 jurisdictions (44% of the Model 3 population) had data across all these control variables.

## DISCUSSION

Our findings demonstrate a clear association between a higher GHS Index (and component category scores) and reduced age-standardised excess mortality through the Covid-19 pandemic (2020–21) for *non-island* jurisdictions. When correcting for potential weaknesses of previously published analyses, we found that this association does not hold across all jurisdictions, contrary to what has been previously reported. The primary finding can be confirmed by visual inspection of Figure 2 (non-islands), where we see that below a GHS Index overall score of 40, almost all jurisdictions exhibit (cube root [∛] transformed) age-standardised excess mortality >5, whereas above a GHS Index score of 60, almost all jurisdictions are <5. We did not find any strong evidence that the GHS Index was also associated with improved economic growth through the first two years of the Covid-19 pandemic.

The potential weaknesses of previously published analyses, which we have attempted to correct, include our use of 2019 GHS Index scores, which are not influenced by pandemic outcomes, rather than 2021 scores. Use of purchasing power parity GDP rather than current value, to account for inflation and the real value of currency. Also, we transformed right-skewed data (namely excess mortality and GDP) to make it more appropriate for linear models and we included some important social and governance variables as controls in our analyses. Each of these analytic choices seems more appropriate than it’s alternative, when attempting to establish if there is a relationship between GHS Index scores and Covid-19 pandemic outcomes. We stepped through these modifications in a series of models to demonstrate the impact these analytic decisions have. As a result we are confident we demonstrate important findings. We have also flagged issues with the data distributions of GHS Index indicator level scores that make analysis of these using linear regression questionable. As such we refrained from such analysis, and hold a healthy scepticism for regression findings in previous analyses at indicator level of the GHS Index.

We have tested the statistical significance of our findings by correcting p-values for multiple comparisons according to a rational analytic approach. Additionally, the primary (non-island, Model 3) finding is statistically significant even when tested against a stringent and conservative Bonferroni corrected p threshold of 0.0009 (that used by Ledesma et al.).^35^ That said, we are very aware that any chosen p-value thresholds are arbitrary and what matters are patterns in the results.^36^

One pattern in our results is abundantly clear. Islands and non-islands are different. The pandemic affected them differently, with islands exhibiting lower mean age-standardised excess mortality. The relationship between GHS Index scores and pandemic outcomes differs between islands and non-islands, and appropriate health security measures and responses to future pandemics are likely to differ between islands and non-islands. In our Model 2 analysis, of GHS Index scores vs age-standardised excess mortality, which controls for GDP alone, the model explained only 1% of the variance in excess mortality for islands (n = 45), whereas the same model explained 45% of the variance for non-islands (n = 137). On the other hand, the sensitivity analysis model (with a smaller analysis population of jurisdictions), which includes all three societal control variables highlighted by Covid NP Collaborators as having a major influence on pandemic outcomes, explained 59% of variance for islands, and 68% for non-islands.

For non-islands, when inspecting results for the GHS Index category scores, we see that the categories Risk Environment, Response, and Health System as most associated with excess mortality (by magnitude of regression coefficient), with Prevention and Detection following in that order. This finding makes intuitive sense. Given that there is a pandemic, then risk environment, response capabilities, and health system capacities are likely to determine outcomes. Whereas prevention and detection capabilities might be supposed more important before a pandemic begins.

For islands, the absence of any meaningful relationship between GHS Index scores and island pandemic outcomes, along with the generally better pandemic outcomes of islands, shows that future iterations of metrics like the GHS Index will need to treat islands differently. The strong protective effect associated with being an island might not appear helpful as geography is not easily modified. However, this attribute can be seen as a proxy for the ability to implement strong border management which in turn supported an exclusion/elimination strategy for responding to the Covid-19 pandemic. Consequently, we propose that the ability to implement an exclusion/elimination strategy in response to severe pandemics is added as a component of a future upgrade of GHS Index design, along with an emphasis on border control components, while recognising the often smaller populations and low-resource settings of many island jurisdictions.

The global health security system operated by the World Health Organization (WHO) includes the International Health Regulations (IHR) which have recently been revised. Negotiations are continuing on a proposed global pandemic agreement.^37^ These regulations and agreements should also reflect the need for countries to manage their borders in the face of future severe pandemics and for the WHO to provide highly strategic global leadership in such situations.^38^

Data coverage issues mean that each additional variable considered reduced the number of jurisdictions available for study, especially with regard to islands, ranging from n = 142 non-islands and n = 47 islands in Model 1 to n = 56 and n = 8 respectively in the sensitivity model. It is possible that there are as yet unapparent systematic differences between jurisdictions for which there is data and for which there is not. It will be important moving forward that data on key societal variables such as government corruption, trust in government, trust in individuals, and other pandemic response related variables be collected from all jurisdictions. These variables are not all correlated. For example, government corruption is correlated with trust in individuals (Pearson’s r = -0.732), but not trust in government (r = -0.030).

Sample size issues notwithstanding, this and previous work,^21^ has demonstrated that characteristics of politics and society appear to show major correlations with pandemic outcomes. The Risk Environment category score had a stronger relationship to Covid-19 mortality than any of the others. This score includes government effectiveness, public confidence in governance, trust in medical and health advice, and related factors. This feature of the GHS Index has previously been noted, suggesting the GHS Index is a good measure of resilience, including preparedness and social capital.^39^ Trust in government and interpersonal trust, along with low levels of corruption have previously shown large statistically significant associations with lower standardised infection rates due to Covid-19, as well as higher vaccination coverage among middle-income and high-income countries.^21^

The harms of the Covid-19 pandemic are ongoing and it seems likely to become an endemic disease.^40^ But there is a serious risk of new biological threats. Pathogens such as Influenza A(H5N1) and Mpox have pandemic potential, there is the risk of novel coronaviruses, and the possibility of bioengineered organisms. Modelling suggests we can expect a ‘Covid-19 magnitude’ pandemic to have a return period of approximately 33–50 years and an 18–26% chance over the next decade.^41^ And recent forecasting work suggested a 1–4% chance of a biological catastrophe killing 10% of the human population by the year 2100.^4^

The world needs to continue to build resilience and mitigation capacities against biological threats. Ongoing work is needed in key areas such as metagenomic surveillance and diagnosis (eg, on aircraft wastewater), improved indoor air quality, and accelerated vaccine development and capacity for rapid mass production of new vaccines. Analysis of the Covid-19 pandemic has provided a wealth of information and now this information needs to inform future iterations of preparedness benchmarks (as described in extensive national pandemic reviews such as those conducted in the UK,^9^ Australia,^10^ and New Zealand^11^).

The GHS Index needs to be reviewed and revised so that it is measuring the capabilities that are most strongly associated with effective pandemic preparedness and management. This should include new ways of analysing the relationship between indicator level scores and outcomes than what has been published to date. Doing that will make it a more valid tool for motivating donor funders and governments to invest in the most important areas and put fewer resources into those that appear less associated with good health and economic outcomes.

### Study strengths and limitations

Key strengths of this study include analytic choices made to overcome weaknesses of previous work. These improvements include important data transformations, using the 2019 GHS Index (which was not influenced by Covid-19 outcomes), GDP based on purchasing power parity, and taking a principled hierarchical, rather than an unstructured, approach to the analysis of what is a hierarchical and internally correlated index. The use of three regression models in turn illustrates the potential problems of some earlier work as changing key assumptions changes some results. Our study added to previous findings by demonstrating the significant explanatory benefit of distinguishing islands.

We had to exclude some potentially relevant island jurisdictions from the analyses because they didn’t have GHS Index scores, including Bermuda, Greenland, Taiwan, Tokelau, American Samoa, Guam, Northern Mariana Islands, Puerto Rico, and the Virgin Islands. Also, some islands have GHS Index data but lack World Bank GDP data eg, Cook Islands, and Niue. We further note that the definition of islands is complex, see Methods.

Model fit in our macroeconomic analyses was poor and GDP has some limitations as a metric. Changes in calculated annual growth rates are affected by the economic performance of the previous year. For example, a high growth rate does not necessarily reflect the strength of the economy over the past year, it might reflect poor economic performance of the previous year. For reasons such as this we used the pre-pandemic 5-year geometric mean of GDP when calculating growth rates. Although we additionally controlled for government corruption, there are more variables of interest. Unfortunately wider data collection is needed so that the majority of jurisdictions can be included in future controlled analyses.

Ultimately, this was a correlation study and we can’t infer causation. However, there are clear and plausible links between the GHS Index indicators and both health and economic outcomes. The Index has previously demonstrated face and external validity, and the Covid-19 pandemic has provided prospective validation. Our findings are sufficiently strong that they support use of the GHS Index (with appropriate modifications for assessing border biosecurity capability) for guiding priorities for investment in future pandemic preparedness.

## CONCLUSION

The GHS Index predicted better health outcomes in terms of age-standardised excess mortality through the first two years (2020–2021) of the Covid-19 pandemic. However, this finding applies to non-island jurisdictions only, with the GHS Index of island jurisdictions showing no similar associations across any models. Analysis of data from the Covid-19 pandemic revealed which GHS Index categories are more and less strongly associated with pandemic outcomes, and this finding should inform future iterations of preparedness indices.

The analysis highlights a striking difference in the health security characteristics of islands and non-islands. This finding suggests that border biosecurity, which island states have by virtue of their geography but other states need to generate by design, needs much greater focus in the GHS Index. These border control capacities and capabilities need to be able to support exclusion/elimination strategies, and anticipation of rapid vaccine roll-out.

The lack of association with economic outcomes provides some evidence that factors other than health security capabilities, national wealth and government corruption drove macroeconomic outcomes during 2020-2021. More research on this economic question would be valuable, as jurisdictions look to prioritise the capacities and capabilities that are more strongly correlated with health outcomes.

## Acknowledgements

The authors thank Prof Austin Schumacher and A/Prof Joseph Dieleman from the GBD Collaboration for responding to requests and sharing GBD data. We are also immensely thankful for thoughtful feedback from an anonymous reviewer which has substantially improved the methodology of this paper.

## Patient or public involvement

It was not appropriate or possible to involve patients or the public in the design, or conduct, or reporting, or dissemination plans of our research

## Data availability

Data (csv), R Code (rtf), and Regression Results (xls): https://adaptresearchwriting.com/wp-content/uploads/2024/12/241219-ghs-index-covid-data_code_results.zip

