## Supplementary Material for "Global Health Security Index is associated with Covid-19 Pandemic Mortality 2020–2021 but not for Island Jurisdictions"

Data (csv), R code (rtf), & raw regression results (xls) available here: <https://adaptresearchwriting.com/wp-content/uploads/2024/12/241219-ghs-index-covid-data_code_results.zip>

### Data Coverage

**Table S1:** Number of jurisdictions with available data for each variable

|  | **All Jurisdictions (n)** | **Non-Islands**  **(n)** | **Islands**  **(n)** |
| --- | --- | --- | --- |
| Age-standardised excess mortality (2020-21) | 204 | 146 | 58 |
| GDP Growth 2020–2021 | 183 | 136 | 47 |
| GHS Index scores (2019, 2021) | 194 | 145 | 49 |
| ***Control variables*** |  |  |  |
| GDP per capita mean 2015–2019 (PPP, 2017) | 184 | 137 | 47 |
| GDP per capita + government corruption | 155 | 128 | 27 |
| GDP per capita + government corruption + trust in government | 105 | 89 | 16 |
| GDP per capita + government corruption + trust in government + trust in individuals | 64 | 56 | 8 |

Most jurisdictions (islands and non-islands) had data describing mortality, GDP and GHS Index scores. But far fewer jurisdictions, notably islands, had available data for the other three potential control variables, except for government corruption, where data existed for 88% of non-islands and 55% of islands that also had GHS Index scores.

### Distributions

**Figure S1**: Histogram showing distribution of baseline 5-year GDP (PPP) per capita (2015-2019), and age-standardised excess mortality (2020–2021), both before (left) and after (right) their respective natural logarithm and cube root transformations

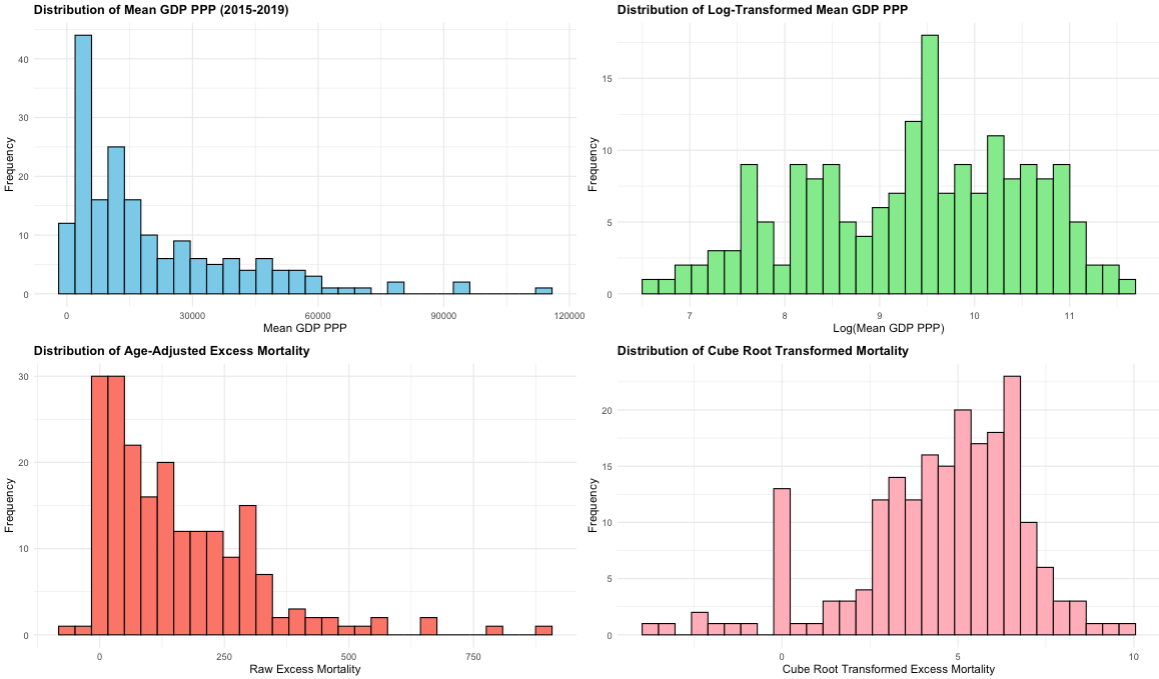

**Figure S2**: Distributions of 2019 GHS Index overall score, and six category scores (untransformed)

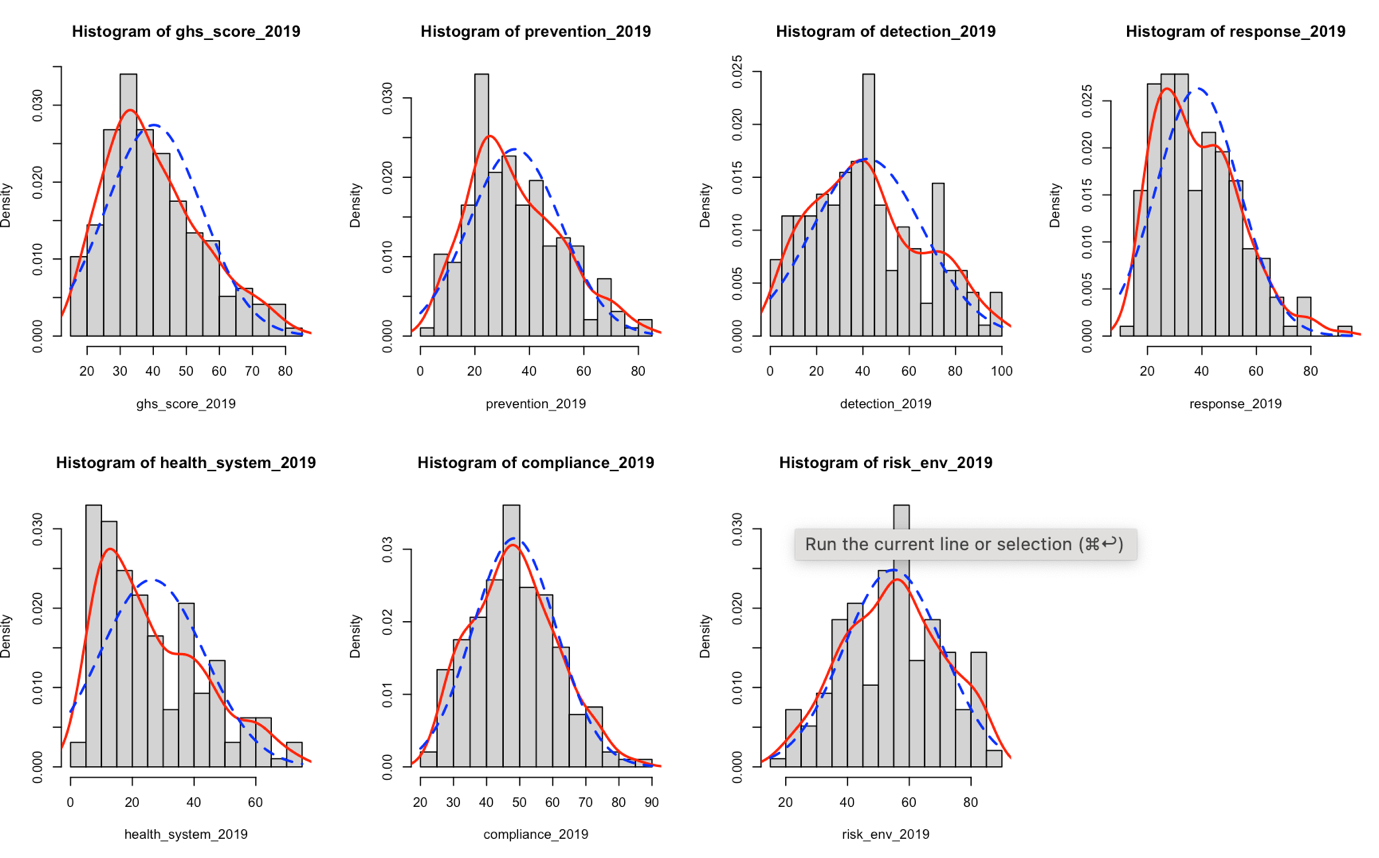

**Figure S3 (a-d)**: Histograms showing how the distributions of the GHS Index indicator level scores violate assumptions of linear regression, which would call into question any conclusions drawn from such analysis.

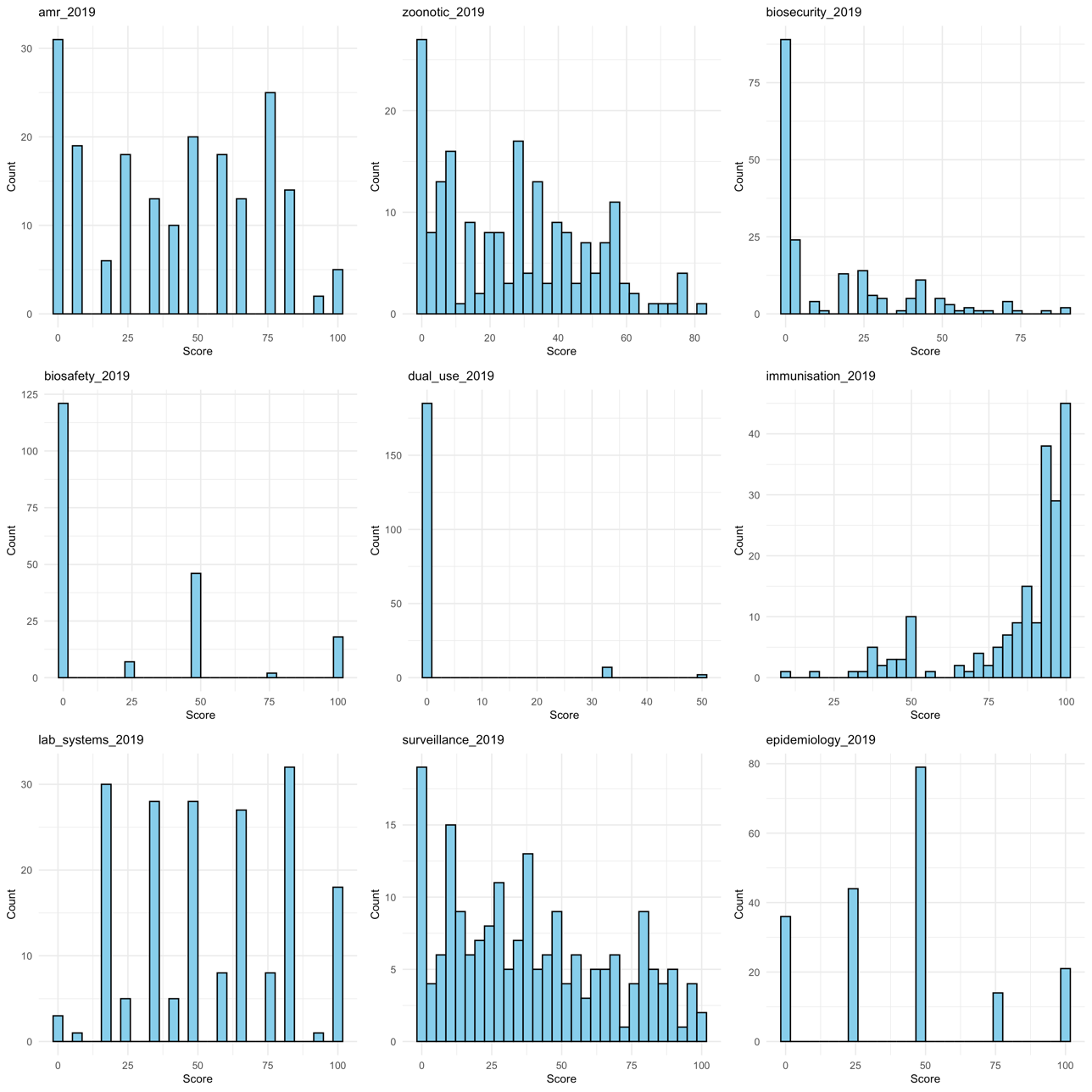

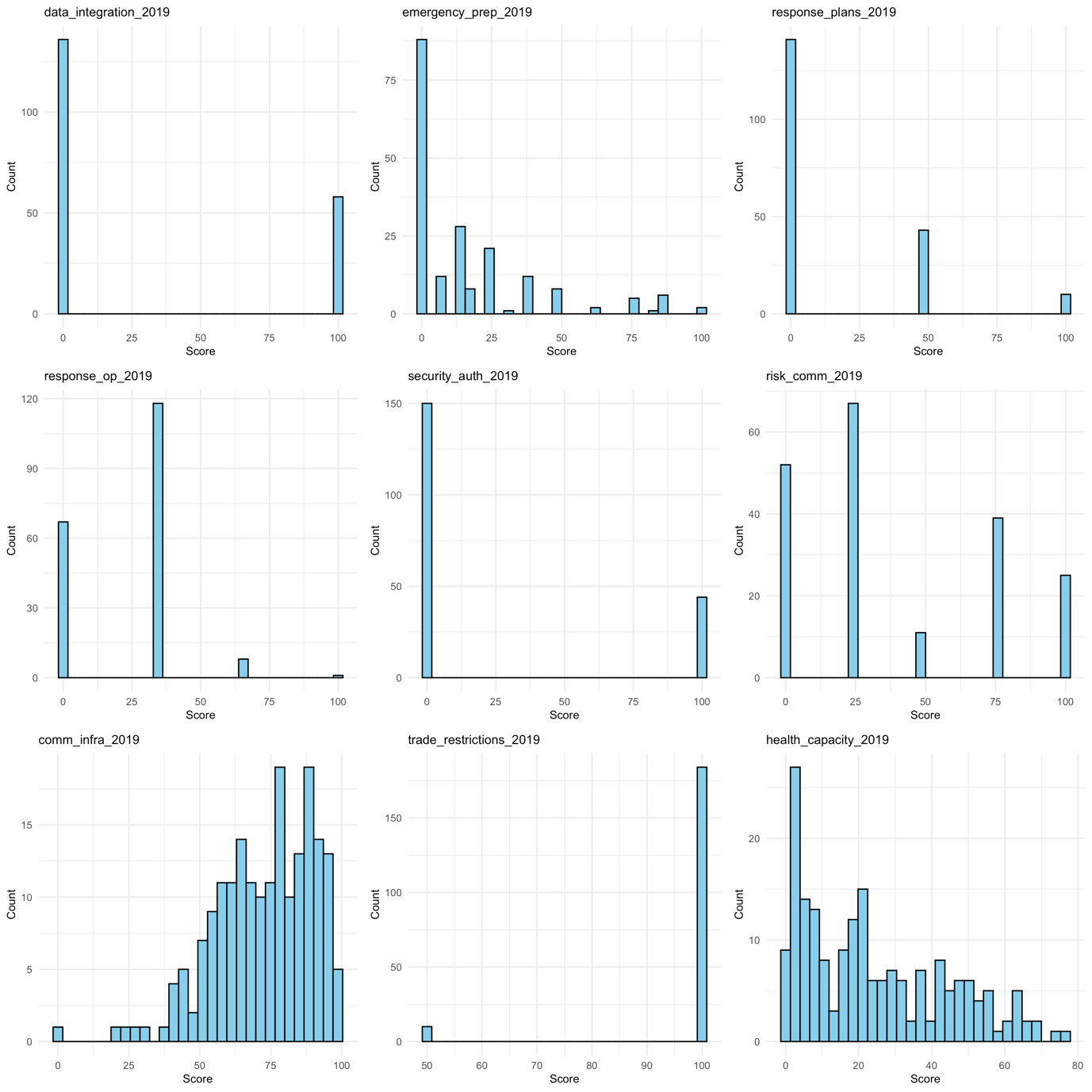

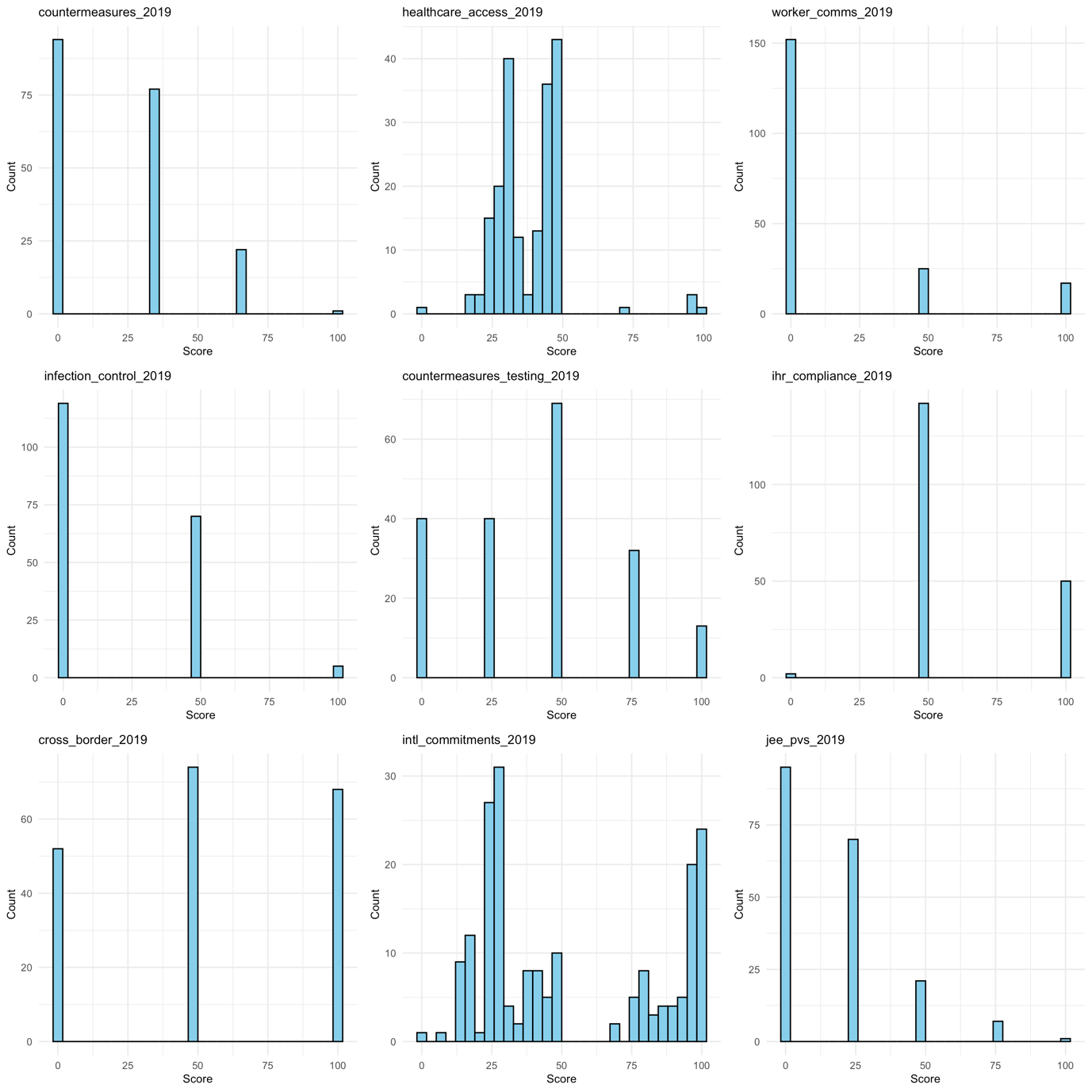

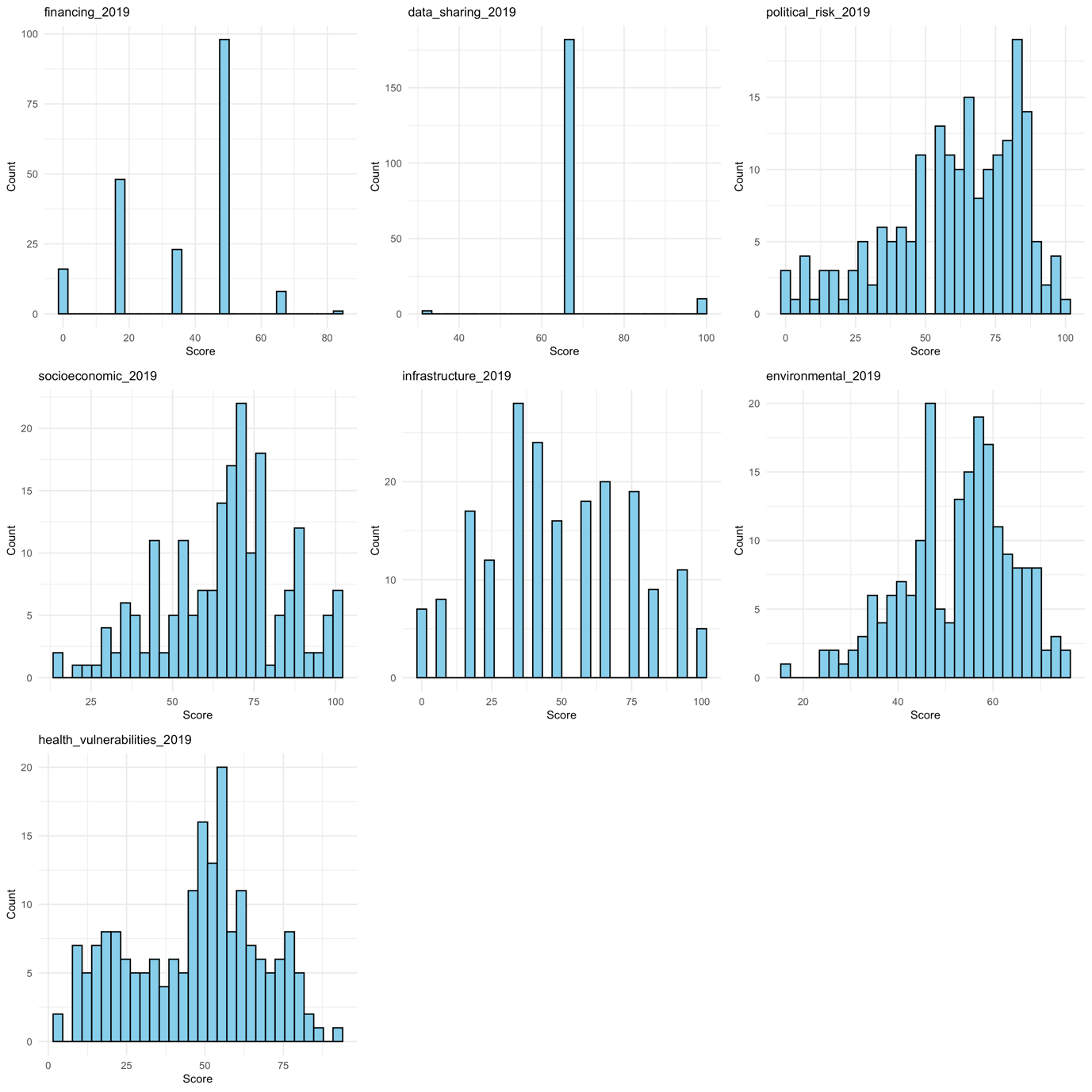

### Regression Equations

For analysis of GHS Index overall score and six category scores only.

#### Model 1 (following Ledesma et al.):

Equation 1.1:

(age standardised excess mortality) = β₀ + β₁(GHS Index 2021) + β₂(GDP current $) + ε

Equation 1.2:

(GDP growth 2019-2020) = β₀ + β₁(GHS Index 2021) + β₂(GDP current $) + ε

Equation 1.3:

(GDP growth 2020-2021) = β₀ + β₁(GHS Index 2021) + β₂(GDP current $) + ε

#### Model 2 (pre-pandemic GHS Index scores with appropriate variable transformations):

Equation 2.1:

∛(age standardised excess mortality) = β₀ + β₁(GHS Index 2019) + β₂.ln(GDP PPP) + ε

Equation 2.2:

(GDP growth 2019-2020) = β₀ + β₁(GHS Index 2019) + β₂.ln(GDP PPP) + ε

Equation 2.3:

(GDP growth 2020-2021) = β₀ + β₁(GHS Index 2019) + β₂.ln(GDP PPP) + ε

#### Model 3 (as Model 2, +controlling for government corruption)

Equation 3.1:

∛(age standardised excess mortality) = β₀ + β₁(GHS Index 2019) + β₂.ln(GDP PPP) + β_3_(government corruption) + ε

Equation 3.2:

(GDP growth 2019-2020) = β₀ + β₁(GHS Index 2019) + β₂.ln(GDP PPP) + β_3_(government corruption) + ε

Equation 3.3:

(GDP growth 2020-2021) = β₀ + β₁(GHS Index 2019) + β₂.ln(GDP PPP) + β_3_(government corruption) + ε

### Regression Results

#### Level 1 of Hierarchical Analysis: GHS Index Overall Score

**Table S2: Age-standardised Excess Mortality 2020-2021 vs GHS Index Overall Score (2021 and 2019) across all analyses**

| **Group** | **Model** | **Dependent Variable** | **Independent Variable** | **Control Variables** | **Coefficient (SE)** | **T-Value** | **P-Value** | **R²** | **Adj. R²** | **N** |
| --- | --- | --- | --- | --- | --- | --- | --- | --- | --- | --- |
| All | Model 1 | EM | GHSI 2021 | GDP | -3.091 (0.86) | -3.61 | 0.000387* | 0.22 | 0.21 | 189 |
| All | Model 2 | cube_r_EM | GHSI 2019 | ln_GDP | 0.011 (0.01) | 0.79 | 0.432050 | 0.20 | 0.19 | 182 |
| All | Model 3 | cube_r_EM | GHSI 2019 | ln_GDP + corruption | -0.005 (0.01) | -0.38 | 0.704835 | 0.46 | 0.45 | 155 |
| All | Sensitivity | cube_r_EM | GHSI 2019 | ln_GDP + corruption + trust govt + trust indivs | -0.006 (0.02) | -0.27 | 0.789155 | 0.60 | 0.57 | 64 |
| Islands | Model 1 | EM | GHSI 2021 | GDP | 0.647 (1.53) | 0.42 | 0.675088 | 0.10 | 0.06 | 47 |
| Islands | Model 2 | cube_r_EM | GHSI 2019 | ln_GDP | 0.027 (0.04) | 0.75 | 0.458671 | 0.06 | 0.01 | 45 |
| Islands | Model 3 | cube_r_EM | GHSI 2019 | ln_GDP + corruption | 0.012 (0.03) | 0.34 | 0.733720 | 0.53 | 0.46 | 27 |
| Islands | Sensitivity | cube_r_EM | GHSI 2019 | ln_GDP + corruption + trust govt + trust indivs | 0.046 (0.09) | 0.49 | 0.674571 | 0.88 | 0.59 | 8 |
| Non-Islands | Model 1 | EM | GHSI 2021 | GDP | -5.353 (0.89) | -5.99 | <0.000001* | 0.36 | 0.35 | 142 |
| Non-Islands | Model 2 | cube_r_EM | GHSI 2019 | ln_GDP | -0.050 (0.01) | -4.46 | 0.000017* | 0.46 | 0.45 | 137 |
| Non-Islands | Model 3 | cube_r_EM | GHSI 2019 | ln_GDP + corruption | -0.046 (0.01) | -3.48 | 0.000683* | 0.49 | 0.48 | 128 |
| Non-Islands | Sensitivity | cube_r_EM | GHSI 2019 | ln_GDP + corruption + trust govt + trust indivs | -0.052 (0.02) | -3.14 | 0.002844 | 0.71 | 0.68 | 56 |

**Table notes**: Coefficient for Model 1 cannot be compared to other models due to untransformed EM and GDP; Asterisk (*) indicates p-value remains statistically significant against the pre-specified Bonferroni corrected p threshold of 0.0056; cube_r: cube root transformed; EM: age-standardised excess mortality; ln: natural logarithm transformed; SE: standard error; Sensitivity: sensitivity analysis only, including all control variables (note the much lower N across the sensitivity analyses – compare to models 1-3 with caution).

**Table S3: GDP Growth 2019-2020 vs GHS Index Overall Score (2021 and 2019) across all analyses**

| **Group** | **Model** | **Dependent Variable** | **Independent Variable** | **Control Variables** | **Coefficient (SE)** | **T-Value** | **P-Value** | **R²** | **Adj. R²** | **N** |
| --- | --- | --- | --- | --- | --- | --- | --- | --- | --- | --- |
| All | Model 1 | GDP Growth 2019–2020 | GHSI 2021 | GDP | 0.076 (0.03) | 2.71 | 0.007405 | 0.04 | 0.03 | 188 |
| All | Model 2 | GDP Growth 2019–2020 | GHSI 2019 | ln_GDP | 0.057 (0.02) | 3.36 | 0.000951* | 0.07 | 0.06 | 182 |
| All | Model 3 | GDP Growth 2019–2020 | GHSI 2019 | ln_GDP + corruption | 0.061 (0.02) | 2.89 | 0.004457* | 0.09 | 0.07 | 155 |
| All | Sensitivity | GDP Growth 2019–2020 | GHSI 2019 | ln_GDP + corruption + trust govt + trust indivs | -0.026 (0.03) | -0.82 | 0.414487 | 0.21 | 0.15 | 64 |
| Islands | Model 1 | GDP Growth 2019–2020 | GHSI 2021 | GDP | -0.047 (0.07) | -0.71 | 0.479910 | 0.01 | -0.03 | 47 |
| Islands | Model 2 | GDP Growth 2019–2020 | GHSI 2019 | ln_GDP | 0.030 (0.04) | 0.84 | 0.403055 | 0.03 | -0.02 | 45 |
| Islands | Model 3 | GDP Growth 2019–2020 | GHSI 2019 | ln_GDP + corruption | 0.048 (0.05) | 0.95 | 0.351436 | 0.05 | -0.08 | 27 |
| Islands | Sensitivity | GDP Growth 2019–2020 | GHSI 2019 | ln_GDP + corruption + trust govt + trust indivs | 0.018 (0.05) | 0.35 | 0.757940 | 0.84 | 0.45 | 8 |
| Non-Islands | Model 1 | GDP Growth 2019–2020 | GHSI 2021 | GDP | 0.106 (0.03) | 3.23 | 0.001563* | 0.07 | 0.06 | 141 |
| Non-Islands | Model 2 | GDP Growth 2019–2020 | GHSI 2019 | ln_GDP | 0.074 (0.02) | 3.46 | 0.000719* | 0.09 | 0.08 | 137 |
| Non-Islands | Model 3 | GDP Growth 2019–2020 | GHSI 2019 | ln_GDP + corruption | 0.060 (0.03) | 2.36 | 0.019763 | 0.11 | 0.08 | 128 |
| Non-Islands | Sensitivity | GDP Growth 2019–2020 | GHSI 2019 | ln_GDP + corruption + trust govt + trust indivs | -0.027 (0.04) | -0.66 | 0.511367 | 0.20 | 0.13 | 56 |

**Table notes**: Asterisk (*) indicates p-value remains statistically significant against the pre-specified Bonferroni corrected p threshold of 0.0056; ln: natural logarithm transformed; SE: standard error; Sensitivity: sensitivity analysis only, including all control variables (note the much lower N across the sensitivity analyses – compare to models 1-3 with caution).

**Table S4: GDP Growth 2020-2021 vs GHS Index Overall Score (2021 and 2019) across all analyses**

| **Group** | **Model** | **Dependent Variable** | **Independent Variable** | **Control Variables** | **Coefficient (SE)** | **T-Value** | **P-Value** | **R²** | **Adj. R²** | **N** |
| --- | --- | --- | --- | --- | --- | --- | --- | --- | --- | --- |
| All | Model 1 | GDP Growth 2020–2021 | GHSI 2021 | GDP | 0.084 (0.03) | 3.29 | 0.001202* | 0.07 | 0.06 | 187 |
| All | Model 2 | GDP Growth 2020–2021 | GHSI 2019 | ln_GDP | 0.060 (0.02) | 3.47 | 0.000649* | 0.07 | 0.06 | 181 |
| All | Model 3 | GDP Growth 2020–2021 | GHSI 2019 | ln_GDP + corruption | 0.053 (0.02) | 2.47 | 0.014449 | 0.06 | 0.04 | 155 |
| All | Sensitivity | GDP Growth 2020–2021 | GHSI 2019 | ln_GDP + corruption + trust govt + trust indivs | -0.019 (0.03) | -0.57 | 0.568738 | 0.09 | 0.01 | 64 |
| Islands | Model 1 | GDP Growth 2020–2021 | GHSI 2021 | GDP | 0.018 (0.07) | 0.24 | 0.810317 | 0.02 | -0.03 | 46 |
| Islands | Model 2 | GDP Growth 2020–2021 | GHSI 2019 | ln_GDP | 0.051 (0.04) | 1.41 | 0.166045 | 0.05 | 0.01 | 45 |
| Islands | Model 3 | GDP Growth 2020–2021 | GHSI 2019 | ln_GDP + corruption | 0.044 (0.05) | 0.89 | 0.382546 | 0.08 | -0.04 | 27 |
| Islands | Sensitivity | GDP Growth 2020–2021 | GHSI 2019 | ln_GDP + corruption + trust govt + trust indivs | 0.036 (0.05) | 0.78 | 0.517816 | 0.81 | 0.33 | 8 |
| Non-Islands | Model 1 | GDP Growth 2020–2021 | GHSI 2021 | GDP | 0.093 (0.03) | 3.20 | 0.001728* | 0.08 | 0.07 | 141 |
| Non-Islands | Model 2 | GDP Growth 2020–2021 | GHSI 2019 | ln_GDP | 0.059 (0.02) | 2.72 | 0.007440 | 0.06 | 0.04 | 136 |
| Non-Islands | Model 3 | GDP Growth 2020–2021 | GHSI 2019 | ln_GDP + corruption | 0.049 (0.03) | 1.89 | 0.060968 | 0.07 | 0.05 | 128 |
| Non-Islands | Sensitivity | GDP Growth 2020–2021 | GHSI 2019 | ln_GDP + corruption + trust govt + trust indivs | -0.027 (0.04) | -0.66 | 0.513563 | 0.09 | 0.00 | 56 |

**Table notes**: Asterisk (*) indicates p-value remains statistically significant against the pre-specified Bonferroni corrected p threshold of 0.0056; ln: natural logarithm transformed; SE: standard error; Sensitivity: sensitivity analysis only, including all control variables (note the much lower N across the sensitivity analyses – compare to models 1-3 with caution).

#### Level 2 of Hierarchical Analysis: GHS Index Category Scores for Significant Level 1 Results

##### Main Regression Result (Model 3)

Model 3 is the improved model using 2019 GHS Index scores as independent variables (because 2021 scores were influenced by the Covid-19 pandemic), with cube root transformed age-standardised excess mortality 2020-21 as dependent variable, natural logarithm transformed GDP (PPP) per capita and government corruption as control variables. Model 3 retains reasonable N, unlike the sensitivity analysis, which includes additional control variables, but at the cost of analysing only a minority of jurisdictions. The analysis excludes GHS Index indicator-level scores due to extreme varying and non-normal distributions (see histograms above).

**Table S5: Age-standardised Excess Mortality 2020–2021 (Model 3)**

| **Group** | **Dependent Variable** | **Independent Variable** | **Control Variables** | **Coefficient (SE)** | **T-Value** | **P-Value** | **BH corrected P-Value** | **R²** | **Adj. R²** | **N** |
| --- | --- | --- | --- | --- | --- | --- | --- | --- | --- | --- |
| All | EM | GHSI Overall | GDP + corruption | -0.005 (0.01) | -0.38 | 0.704835 |  | 0.46 | 0.45 | 155 |
| All | EM | Prevention | GDP + corruption | 0.003 (0.01) | 0.32 | 0.749319 |  | 0.46 | 0.45 | 155 |
| All | EM | Detection | GDP + corruption | -0.002 (0.01) | -0.31 | 0.756928 |  | 0.46 | 0.45 | 155 |
| All | EM | Response | GDP + corruption | -0.005 (0.01) | -0.46 | 0.645192 |  | 0.46 | 0.45 | 155 |
| All | EM | Health System | GDP + corruption | 0.000 (0.01) | 0.03 | 0.974923 |  | 0.46 | 0.45 | 155 |
| All | EM | Compliance | GDP + corruption | -0.005 (0.01) | -0.40 | 0.692028 |  | 0.46 | 0.45 | 155 |
| All | EM | Risk Environment | GDP + corruption | -0.057 (0.03) | -2.28 | 0.023892 |  | 0.47 | 0.46 | 155 |
| Islands | EM | GHSI Overall | GDP + corruption | 0.012 (0.03) | 0.34 | 0.733720 |  | 0.53 | 0.46 | 27 |
| Islands | EM | Prevention | GDP + corruption | 0.006 (0.03) | 0.21 | 0.835565 |  | 0.52 | 0.46 | 27 |
| Islands | EM | Detection | GDP + corruption | 0.006 (0.02) | 0.32 | 0.748827 |  | 0.53 | 0.46 | 27 |
| Islands | EM | Response | GDP + corruption | 0.024 (0.02) | 0.98 | 0.337617 |  | 0.54 | 0.48 | 27 |
| Islands | EM | Health System | GDP + corruption | 0.003 (0.03) | 0.10 | 0.920240 |  | 0.52 | 0.46 | 27 |
| Islands | EM | Compliance | GDP + corruption | 0.000 (0.03) | 0.00 | 0.997709 |  | 0.52 | 0.46 | 27 |
| Islands | EM | Risk Environment | GDP + corruption | -0.147 (0.09) | -1.63 | 0.115768 |  | 0.57 | 0.52 | 27 |
| **Non-Islands** | EM | GHSI Overall | GDP + corruption | -0.046 (0.01) | -3.48 | 0.000683* |  | 0.49 | 0.48 | 128 |
| **Non-Islands** | EM | Prevention | GDP + corruption | -0.024 (0.01) | -2.20 | **0.029701** | **0.044551** | 0.46 | 0.45 | 128 |
| **Non-Islands** | EM | Detection | GDP + corruption | -0.019 (0.01) | -2.99 | **0.003327** | **0.014004** | 0.48 | 0.47 | 128 |
| **Non-Islands** | EM | Response | GDP + corruption | -0.029 (0.01) | -2.88 | **0.004668** | **0.014004** | 0.48 | 0.46 | 128 |
| **Non-Islands** | EM | Health System | GDP + corruption | -0.027 (0.01) | -2.57 | **0.011423** | **0.022846** | 0.47 | 0.46 | 128 |
| **Non-Islands** | EM | Compliance | GDP + corruption | -0.018 (0.01) | -1.60 | 0.113249 | 0.135899 | 0.45 | 0.44 | 128 |
| **Non-Islands** | EM | Risk Environment | GDP + corruption | -0.058 (0.02) | -2.57 | **0.011248** | **0.022846** | 0.47 | 0.46 | 128 |

**Table note:** Asterisk (*) represents statistical significance after Bonferroni correction in the primary analysis (GHS Index overall score, see Table S2 above)**.** Therefore, the non-islands group in Model 3 was further analysed to correct p-values with Benjamini-Hochberg (BH) correction (noting 12 comparisons between category variables and dependent variables, 6 for excess mortality, and 6 for GDP growth 2019-20, see Table S6 below). Bold represents statistical significance after BH correction with false discovery rate 0.05.

**Table S6: GDP Growth 2019–2020 (Model 3)**

| **Group** | **Dependent Variable** | **Independent Variable** | **Control Variables** | **Coefficient (SE)** | **T-Value** | **P-Value** | **BH corrected P-Value** | **R²** | **Adj. R²** | **N** |
| --- | --- | --- | --- | --- | --- | --- | --- | --- | --- | --- |
| **All** | GDP Growth 2019-20 | 2019 GHSI Overall Score | GDP + corrupt | 0.061 (0.02) | 2.89 | 0.004457* |  | 0.09 | 0.07 | 155 |
| **All** | GDP Growth 2019-20 | Prevention | GDP + corrupt | 0.058 (0.02) | 3.42 | **0.000803** | **0.009633** | 0.11 | 0.09 | 155 |
| **All** | GDP Growth 2019-20 | Detection | GDP + corrupt | 0.031 (0.01) | 2.91 | **0.004151** | **0.014004** | 0.09 | 0.07 | 155 |
| **All** | GDP Growth 2019-20 | Response | GDP + corrupt | 0.017 (0.02) | 0.99 | 0.321900 | 0.321900 | 0.04 | 0.02 | 155 |
| **All** | GDP Growth 2019-20 | Health System | GDP + corrupt | 0.038 (0.02) | 2.20 | **0.029654** | **0.044551** | 0.07 | 0.05 | 155 |
| **All** | GDP Growth 2019-20 | Compliance | GDP + corrupt | 0.031 (0.02) | 1.63 | 0.106071 | 0.135899 | 0.05 | 0.03 | 155 |
| **All** | GDP Growth 2019-20 | Risk Environment | GDP + corrupt | 0.058 (0.04) | 1.39 | 0.165136 | 0.180148 | 0.05 | 0.03 | 155 |
| Islands | GDP Growth 2019-20 | 2019 GHSI Overall Score | GDP + corrupt | 0.048 (0.05) | 0.95 | 0.351436 |  | 0.05 | -0.08 | 27 |
| Islands | GDP Growth 2019-20 | Prevention | GDP + corrupt | 0.057 (0.04) | 1.33 | 0.195626 |  | 0.08 | -0.04 | 27 |
| Islands | GDP Growth 2019-20 | Detection | GDP + corrupt | 0.045 (0.03) | 1.66 | 0.110118 |  | 0.11 | 0.00 | 27 |
| Islands | GDP Growth 2019-20 | Response | GDP + corrupt | -0.003 (0.04) | -0.07 | 0.943287 |  | 0.01 | -0.12 | 27 |
| Islands | GDP Growth 2019-20 | Health System | GDP + corrupt | 0.050 (0.05) | 1.10 | 0.281720 |  | 0.06 | -0.07 | 27 |
| Islands | GDP Growth 2019-20 | Compliance | GDP + corrupt | -0.007 (0.05) | -0.15 | 0.884932 |  | 0.01 | -0.12 | 27 |
| Islands | GDP Growth 2019-20 | Risk Environment | GDP + corrupt | -0.010 (0.14) | -0.07 | 0.945380 |  | 0.01 | -0.12 | 27 |
| Non-Islands | GDP Growth 2019-20 | 2019 GHSI Overall Score | GDP + corrupt | 0.060 (0.03) | 2.36 | 0.019763 |  | 0.11 | 0.08 | 128 |
| Non-Islands | GDP Growth 2019-20 | Prevention | GDP + corrupt | 0.056 (0.02) | 2.78 | 0.006319 |  | 0.12 | 0.10 | 128 |
| Non-Islands | GDP Growth 2019-20 | Detection | GDP + corrupt | 0.024 (0.01) | 1.93 | 0.056191 |  | 0.09 | 0.07 | 128 |
| Non-Islands | GDP Growth 2019-20 | Response | GDP + corrupt | 0.018 (0.02) | 0.93 | 0.354557 |  | 0.07 | 0.05 | 128 |
| Non-Islands | GDP Growth 2019-20 | Health System | GDP + corrupt | 0.030 (0.02) | 1.50 | 0.135894 |  | 0.08 | 0.06 | 128 |
| Non-Islands | GDP Growth 2019-20 | Compliance | GDP + corrupt | 0.038 (0.02) | 1.78 | 0.076977 |  | 0.09 | 0.07 | 128 |
| Non-Islands | GDP Growth 2019-20 | Risk Environment | GDP + corrupt | 0.064 (0.04) | 1.47 | 0.143407 |  | 0.08 | 0.06 | 128 |

**Table note**: poor model fit based on adjusted R^2^; asterisk (*) represents statistical significance after Bonferroni correction in the primary analysis (GHS Index overall score, see Table S3 above)**.** Therefore, the ‘all jurisdictions’ group in Model 3 was further analysed to correct p-values with Benjamini-Hochberg (BH) correction (noting 12 comparisons between category variables and dependent variables, 6 for excess mortality [above], and 6 for GDP growth 2019-20 [this table]). Bold represents statistical significance after BH correction with false discovery rate 0.05.

**Table S7: GDP Growth 2020-2021**

| **Group** | **Dependent Variable** | **Independent Variable** | **Control Variables** | **Coefficient (SE)** | **T-Value** | **P-Value** | **R²** | **Adj. R²** | **N** |
| --- | --- | --- | --- | --- | --- | --- | --- | --- | --- |
| All | GDP Growth 2020-21 | 2019 GHSI Overall Score | GDP + corruption | 0.053 (0.02) | 2.47 | 0.014449 | 0.06 | 0.04 | 155 |
| All | GDP Growth 2020-21 | Prevention | GDP + corruption | 0.054 (0.02) | 3.13 | 0.002090 | 0.08 | 0.06 | 155 |
| All | GDP Growth 2020-21 | Detection | GDP + corruption | 0.027 (0.01) | 2.43 | 0.016115 | 0.06 | 0.04 | 155 |
| All | GDP Growth 2020-21 | Response | GDP + corruption | 0.015 (0.02) | 0.88 | 0.378361 | 0.03 | 0.01 | 155 |
| All | GDP Growth 2020-21 | Health System | GDP + corruption | 0.032 (0.02) | 1.84 | 0.067479 | 0.04 | 0.02 | 155 |
| All | GDP Growth 2020-21 | Compliance | GDP + corruption | 0.021 (0.02) | 1.07 | 0.284305 | 0.03 | 0.01 | 155 |
| All | GDP Growth 2020-21 | Risk Environment | GDP + corruption | 0.077 (0.04) | 1.83 | 0.068838 | 0.04 | 0.02 | 155 |
| Islands | GDP Growth 2020-21 | 2019 GHSI Overall Score | GDP + corruption | 0.044 (0.05) | 0.89 | 0.382546 | 0.08 | -0.04 | 27 |
| Islands | GDP Growth 2020-21 | Prevention | GDP + corruption | 0.050 (0.04) | 1.19 | 0.246053 | 0.10 | -0.02 | 27 |
| Islands | GDP Growth 2020-21 | Detection | GDP + corruption | 0.038 (0.03) | 1.44 | 0.163045 | 0.13 | 0.01 | 27 |
| Islands | GDP Growth 2020-21 | Response | GDP + corruption | 0.009 (0.04) | 0.25 | 0.805769 | 0.05 | -0.08 | 27 |
| Islands | GDP Growth 2020-21 | Health System | GDP + corruption | 0.037 (0.04) | 0.84 | 0.409448 | 0.07 | -0.05 | 27 |
| Islands | GDP Growth 2020-21 | Compliance | GDP + corruption | -0.005 (0.05) | -0.11 | 0.911280 | 0.05 | -0.08 | 27 |
| Islands | GDP Growth 2020-21 | Risk Environment | GDP + corruption | -0.041 (0.14) | -0.29 | 0.772750 | 0.05 | -0.07 | 27 |
| Non-Islands | GDP Growth 2020-21 | 2019 GHSI Overall Score | GDP + corruption | 0.049 (0.03) | 1.89 | 0.060968 | 0.07 | 0.05 | 128 |
| Non-Islands | GDP Growth 2020-21 | Prevention | GDP + corruption | 0.051 (0.02) | 2.51 | 0.013524 | 0.09 | 0.07 | 128 |
| Non-Islands | GDP Growth 2020-21 | Detection | GDP + corruption | 0.019 (0.01) | 1.50 | 0.136682 | 0.06 | 0.04 | 128 |
| Non-Islands | GDP Growth 2020-21 | Response | GDP + corruption | 0.011 (0.02) | 0.57 | 0.566648 | 0.04 | 0.02 | 128 |
| Non-Islands | GDP Growth 2020-21 | Health System | GDP + corruption | 0.025 (0.02) | 1.23 | 0.222497 | 0.05 | 0.03 | 128 |
| Non-Islands | GDP Growth 2020-21 | Compliance | GDP + corruption | 0.023 (0.02) | 1.09 | 0.276751 | 0.05 | 0.03 | 128 |
| Non-Islands | GDP Growth 2020-21 | Risk Environment | GDP + corruption | 0.090 (0.04) | 2.07 | 0.040728 | 0.07 | 0.05 | 128 |

**Table note**: poor model fit based on adjusted R^2^
